# Motor Learning in Response to Different Experimental Pain Models among Healthy Individuals: A Systematic Review

**DOI:** 10.1101/2021.08.24.21262581

**Authors:** Mohammad Izadi, Sae Franklin, Marianna Bellafiore, David W. Franklin

## Abstract

Learning new movement patterns is a normal part of daily life, but of critical importance in both sport and rehabilitation. A major question is how different sensory signals are integrated together to give rise to motor adaptation and learning. More specifically, there is growing evidence that pain can give rise to alterations in the learning process. Despite a number of studies investigating the role of pain on the learning process, there is still no systematic review to summarize and critically assess investigations regarding this topic in the literature. Here in this systematic review, we summarize and critically evaluate studies that examined the influence of experimental pain on motor learning. Seventeen studies that exclusively assessed the effect of experimental pain models on motor learning among healthy individuals were included for this systematic review, carried out based on the PRISMA statement. The results of the review revealed there is no consensus regarding the effect of pain on the skill learning acquisition and retention. However, several studies demonstrated that participants who experienced pain continued to express a changed motor strategy to perform a motor task even one week after training under the pain condition. The results highlight a need for further studies in this area of research, and specifically to investigate whether pain has different effects on motor learning depending on the type of motor task.

## Introduction

Pain is an unpleasant but important perception, in order to attract attention and avoid further damage to the body. However, patients and athletes are often required to learn new movement patterns as part of a rehabilitation program in the presence of pain conditions. While it may be necessary to perform rehabilitation exercise immediately after an injury in order to return to optimal performance, there is concern surrounding the effect of pain on the learning process. That is, it has been reported that pain, as a sensory input, might affect the sensorimotor system leading to changes in motor performance, including redistribution of muscle activation patterns, and a reduction in muscle endurance that is essential for performing dynamic motor skills (Bank et al., 2013; Hodges & Tucker, 2011; Lamothe et al., 2014). In this vein, several studies (Dancey et al., 2014; Dancey et al., 2016b; Mavromatis et al., 2017) demonstrated that pain can give rise to neuroplastic changes in the cortex. However, these neuroplastic changes have been associated with both decreases in motor performance (Mavromatis et al., 2017) and improvements in motor learning outcomes in response to pain (Dancey et al., 2016a).

In order to investigate the influence of pain on the learning process, experimental pain models, including muscle and cutaneous pain, have been used to test its effect on motor adaptation (Dancey et al., 2016a; Dancey et al., 2014; Dancey et al., 2019; Lamothe et al., 2014). Other studies have examined the impact of chronic pain on motor learning (Vallence et al., 2013; Zaman et al., 2015). However, chronic pain cannot detect the pure influence of pain on this process, as chronic pain can also be associated with pain-related fear or tissue damage both of which could affect motor learning (Bank et al., 2013). Therefore, using only experimental pain models can assist in studying the pure effect of pain on the learning process.

To date, although many studies (Bilodeau et al., 2016; Bouffard et al., 2014; Dancey et al., 2016a; Dancey et al., 2014; Dancey et al., 2016b; Dancey et al., 2019; Lamothe et al., 2014; Rittig-Rasmussen et al., 2014) have examined the effect of experimental pain models on motor learning; they have provided contradictory findings. For example, some studies have suggested that acute cutaneous pain models improve motor learning acquisition (Dancey et al., 2014; Dancey et al., 2016b) and retention (Dancey et al., 2016b), whereas other studies applying a similar experimental pain model demonstrated no alteration in the learning process (Bilodeau et al., 2016; Bouffard et al., 2014; Rittig-Rasmussen et al., 2014).

Despite the growing literature on the knowledge of the learning process in the presence of experimental pain models, there has been no systematic study reviewing this literature. Considering the contradictory results related to motor learning during pain and the absence of a systemic review, it is important to synthesize and critically assess the studies on motor learning to assess experimental pain models. This information will help to a better understanding of the effect of pain on skill learning acquisition and retention, which is important for developing sport training and rehabilitation programs. Hence, the aim of the current study is to systematically review the research outputs that have examined the effect of experimental pain models on motor learning among healthy individuals.

## Methods

This systematic review was reported based on the Preferred Reporting Items for Systematic Reviews and Meta-Analyses (PRISMA) guidelines (Shamseer et al., 2015).

### Search Strategy

Electronic databases (PubMed, Web of Science, and Embase) were used to search the literature up to April 2021. A combination of free-text terms and MeSH terms regarding motor learning (including retention) and experimental pain was applied (see supplementary material section). Search strategies of relevant systematic reviews were also checked in order to carry out an elaborate strategy. In addition, references of included studies were hand-searched to detect all pertinent studies, as well as the citations of the included studies were checked via Google Scholar.

### Eligibility Criteria

All studies that have the following criteria were included in this systematic review: (1) results of research from healthy human subjects; (2) experimental pain was induced in order to detect the effect of pain on the learning process; (3) original research with full text written in the English language; and (4) all study designs other than all types of reviews, meta-analysis, and letter to editors. Studies that induced pain that can result in structural tissue damage, including pain with eccentric exercise and ischemia, were excluded from this study.

### Study Selection

Extraction of studies was performed by one reviewer, after which two authors independently reviewed retrieved titles and abstracts after removing duplicates. Full-text was also reviewed by the two reviewers to ensure that studies were selected in accordance with the inclusion and exclusion criteria. In the case of disagreement between the two authors surrounding the inclusion or exclusion of a study, the issue was resolved through consultations with a third reviewer.

### Data Collection

In order to collect data, a standard form was used so that the following information was included: (1) pain characteristics (location, type, and intensity); (2) outcome variables (i.e. parameters that were used to assess learning); (3) test protocol; (4) general information about characteristics of subjects; and (5) main results. One author gathered the mentioned data from all included studies and another author checked the collected data to decrease error and bias in data collection.

### Quality Assessment

Two authors independently assessed the quality and bias of all included studies based on a modified version of the checklist for measuring the quality of RCTs and non-RCTs written by Downs and Black (Chuter et al., 2014; Downs & Black, 1998). In the modified version of the checklist, item 27 (power) was changed from 0-5 to 0 or 1 so that a study was scored 1 if the study reported a statistical power ≥ 80%; otherwise, it received 0 (Chuter et al., 2014), so that the overall score of the checklist changed from 32 to 28. The quality of included studies was divided into the following four levels: excellent (26-28), good (20-25), fair (15-19), and poor (≤14) (Pas et al., 2015). Inter-rater reliability for the qualitative items was also measured by using the Kappa correlation coefficient between the two reviewers.

## Results

### Study identification

A total of 3500 articles were generated via electronic databases. The titles and abstracts of 3075 studies were screened after removing 425 duplicates. The full text of 26 studies were assessed in agreement with the inclusion and exclusion criteria in which only 16 studies (Arieh et al., 2021; Bilodeau et al., 2016; Boudreau et al., 2010; Boudreau et al., 2007; Bouffard et al., 2014, 2016; Bouffard et al., 2018; Dancey et al., 2016a; Dancey et al., 2014; Dancey et al., 2016b; Dancey et al., 2018, 2019; Gallina et al., 2018; Ingham et al., 2011; Lamothe et al., 2014; Mavromatis et al., 2017) were included in this review. Finally, one study (Salomoni et al., 2019) was added through a hand-searching of the citations of the relevant studies through Google Scholar; thus, 17 studies were included in this review (Figure 1).

**Figure 1.**
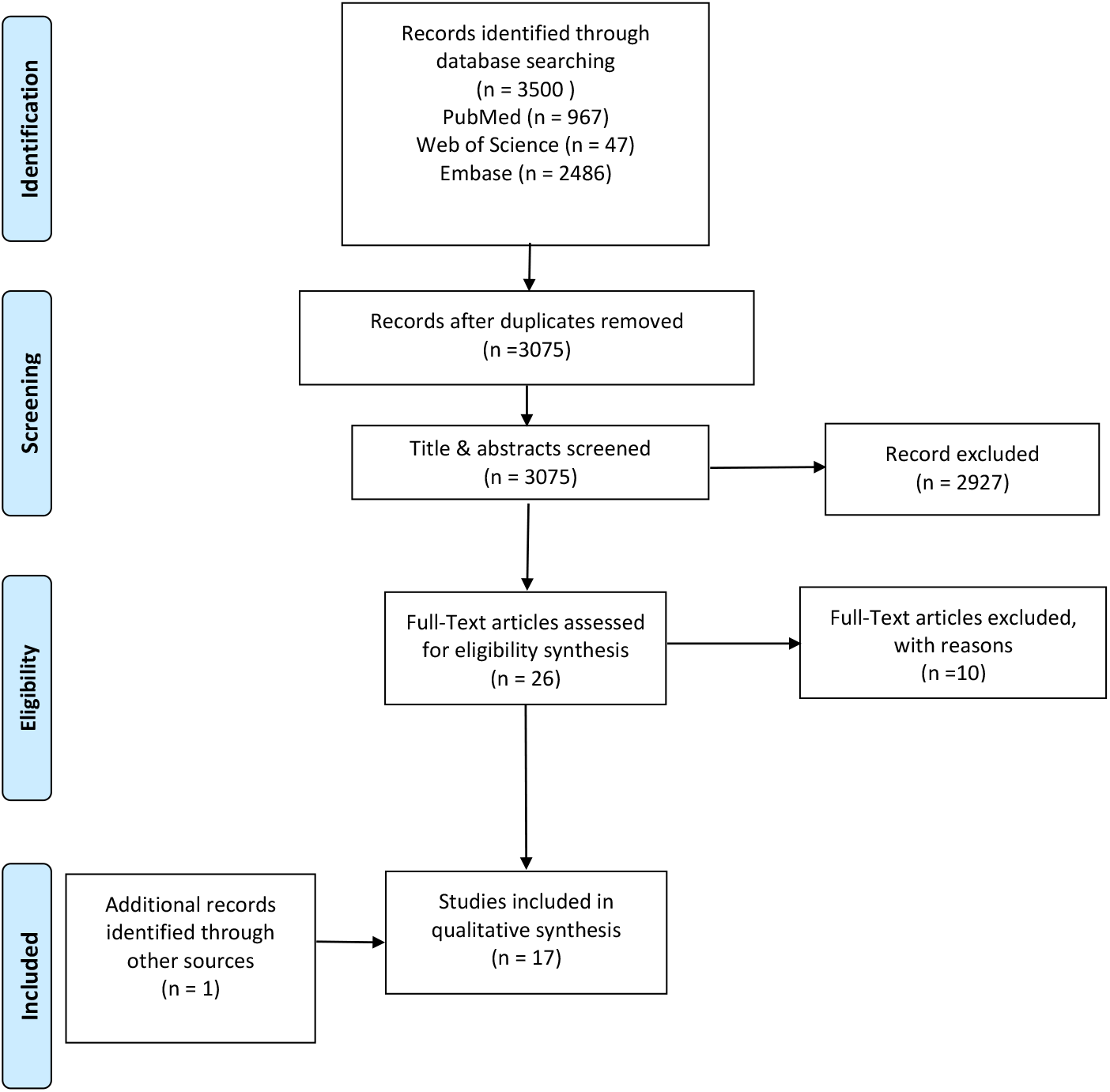
Flow chart of study selection process

### Study characteristics

A total of 484 healthy participants were included across all studies with the ages ranging between 18 and 47 years. Out of the seventeen studies, Arieh and colleagues (Arieh et al., 2021) included only male participants, but both male and female participants were included in the other sixteen studies (Bilodeau et al., 2016; Boudreau et al., 2010; Boudreau et al., 2007; Bouffard et al., 2014, 2016; Bouffard et al., 2018; Dancey et al., 2016a; Dancey et al., 2014; Dancey et al., 2016b; Dancey et al., 2018, 2019; Gallina et al., 2018; Ingham et al., 2011; Lamothe et al., 2014; Mavromatis et al., 2017; Salomoni et al., 2019). Extensive information concerning each study is represented in Table 1.

**Table 1.** study characteristics

| Authors (yrs) | Participants | Pain Characteristics | Testing Procedure | Outcome Measures | Main Results |
| --- | --- | --- | --- | --- | --- |
| Arieh et al (2021) | Healthy (N=30, m) (remote pain, local pain, and control group, N=10), age: 18-25 yrs | Capsaicin gel to the outer side of the elbow 5 cm (local pain); capsaicin gel to the upper part of the knee joint (remote pain); severity of pain was 7- measuring by VAS | Dart-throwing skill during acquisition (with pain) and retention (without pain) ( 1 h, 24 h, and 1 w) phases | Coordination variability pattern during throwing in both acquisition and retention phases (maximum wrist flexion range, maximum elbow extension range, shoulder angular displacement range, angular throw velocity, and throw duration) | No sig effect of pain on dart-throwing learning |
| Bilodeau et al (2016) | Healthy (N=45) (remote pain N=15, f=10, age: 28.5 ± 9.5 yrs); (local pain N=15, f=10, age: 27.4 ± 7.1 yrs); and (control group N=15, f=10, age: 28.8 ± 8.8 yrs) | Thermode 3×3 cm (heat pain) before acquisition phase to the dorsal part of the left wrist (local pain) and the external part of the left leg below the knee joint (remote pain); severity of pain was measured- measuring by NPRS | Finger-tapping task (reproducing the sequence 4-1-3-2-4) during 30s | Error rate and speed of tapping sequences during baseline, post-immediate, post-60 min, and post-24 h (retention) | No sig effect of tonic pain on the acquisition and retention of finger-tapping task |
| Bouffard et al (2014) | Healthy (N=30) (pain group N=15, m=7, age: 26.0 ± 1.4 yrs); (control group N=15, m=8, age: 26.1 ± 2.1 yrs) | Capsaicin gel around the ankle prior to acquisition phase; severity of pain was moderate- measuring by NPRS | Walking task in the presence of a force field adaptation paradigm in two days (acquisition (baseline 1, baseline2, adaptation, and wash-out) and retention (baseline, adaptation, and wash-out)) | A movement error signal that was made based on the ankle angular displacement | Sig effect of tonic pain on the retention phase of a locomotor task, while no sig change in the acquisition phase of gait |
| Bouffard et al (2016) | Healthy (N=37) (pain group N=13, f=8, age: 26.1 ± 1.15 yrs); (control group N=24, f=10, age: 25.8 ± 0.85 yrs) | Capsaicin gel around the ankle between Baseline 1 and Baseline 2 in the first day and prior to baseline in the second day; severity of pain was 5.6 ± 0.7 in Day 1 and 5.5 ± 0.7 in Day 2- measuring by NPRS | Walking task in the presence of a force field adaptation paradigm in two days (acquisition (baseline 1, baseline2, adaptation, and wash-out) and retention (baseline, adaptation, and wash-out)) | A mean absolute error, which was created based on the ankle kinematics, and tibialis anterior ratios that showed TA muscle activation in the adaptation phase relative to baseline | No sig effect of cutaneous pain on total motor performance during both acquisition and retention phases |
| Bouffard et al (2018) | Healthy (N=47) (pain group N=17, f=7, age: 25 ± 1 yrs); (control group N=30, f=14, age: 25 ± 1 yrs) | Hypertonic saline to the tibialis anterior muscle prior to baseline 1 in the first day; the intensity of pain was 5.3 ± 1.2 out of 10- measuring by NPRS | Walking task in the presence of a force field adaptation paradigm in two days (acquisition (baseline 1, baseline2, adaptation, and wash-out) and retention (baseline, adaptation, and wash-out)) | A mean absolute error, which was created based on the ankle kinematics; relative timing of ankle error; tibialis anterior ratios that showed TA muscle activation in the adaptation phase relative to baseline | No sig effect of muscle pain on total motor performance during both acquisition and retention phases |
| Dancey et al (2016) | Healthy (N=24) (pain group N=12, f=8, age: 20.8 ± 3.3 yrs); (control group | Capsaicin gel for pain group and topical cream for the control group in the lateral part of the right elbow; pain intensity average was above | Tracing sequences of sinusoidal pattern waves with various amplitudes and frequencies using | Motor error which showed the average distance of subjects' effort trace from | An improvement in motor learning in response to |
|  | N=12, f=6, age: 22.8 ± 2 yrs) | 4- measuring by NPRS | the thumb in four phases (pre-acquisition, acquisition, post-acquisition, and retention (24-48 h later)) | the displayed sinusoidal wave | cutaneous pain |
| Dancey et al (2016) | Healthy (N=48) ( experiment 1 (N=24; pain group N=12, f=7, age: 20.8 ± 3.3 yrs; control group N=12, f=6, age: 22.8 ± 2 yrs) ( experiment 2 (N=24; remote pain group N=12, f=7, age: 21.8 ± 3.3 yrs; local pain group N=12, f=7, age: 22.9 ± 4.3 yrs) | Capsaicin gel for pain group and topical cream for control group, remote pain and control pain in the lateral part of the dominant elbow and local pain in the Abductor Pollicis Brevis muscle area; pain intensity average was approximately 6 during post-motor learning- measuring by NRPS | A repetitive typing task | Response time and accuracy during a typing task at the begging and end of the motor acquisition and 48 h later (motor learning retention) | An improvement in motor learning retention in the presence of local pain; improved motor performance in the baseline in the presence of acute pain |
| Dancey et al (2018) | Healthy (N=36) (local pain group N=12, f=8, age: 21.2 ± 2.2 yrs); (remote pain group N=12, f=8, age: 20.3 ± 2.5 yrs); (contralateral pain group N=12, f=8, age: 21.4 ± 2.4 yrs) | Capsaicin gel for pain group in which remote and contralateral pain in the lateral part of the dominant and non-dominant elbow, respectively; local pain in the Abductor Pollicis Brevis muscle area; pain intensity average was evaluated- measuring by NPRS | Tracing sequences of sinusoidal pattern waves with various amplitudes and frequencies using thumb in four phases (pre-acquisition, acquisition, post-acquisition, and retention (24-48 h later)) | Motor error which showed the average distance of subjects' effort trace from the displayed sinusoidal wave | No sig effect of pain location on motor learning acquisition and retention |
| Dancey et al (2019) | Healthy (N=24) (pain group N=12, f=9, age: 19.9 ± 0.9 yrs); (control group N=12, f=9, age: 20.7 ± 1.4 yrs) | Capsaicin gel for pain group and topical cream for control group in the lateral part of the dominant elbow; pain intensity average was above 4- measuring by NPRS | Tracing sequences of sinusoidal pattern waves with various amplitudes and frequencies using the thumb in four phases (pre-acquisition, acquisition, post-acquisition, and retention (24-48 h later)) | Mean motor error which showed the average distance of subjects' effort trace from the displayed sinusoidal wave | An improvement in motor performance in the presence of tonic pain |
| Lamothe et al (2014) | Healthy (N=29) (pain group N=15, f=7, age: 25.8 ± 4.1 yrs); (control group N=14, f=8, age: 26.6 ± 4.8 yrs) | Capsaicin gel for pain group above the elbow between two baseline in the first day; pain intensity average was 7.8 ± 0.9 at the initiation of baseline 2- measuring by NPRS | A reaching task in the presence of force field adaptation in four phases (baseline1, baseline2, acquisition, and retention (24h)) | Final error and the initial range of deviation | No sig effect of tonic pain on baseline reaching performance; a larger final error in the pain group than the control group during both acquisition and retention |
| Mavromatis et al (2017) | Healthy (N=30) (pain group N=15, f=6, age: 26 ± 6 yrs); (control group N=15, f=6, age: 27 ± 6 yrs) | Capsaicin gel for pain group around the lateral part of the first metacarpal after the first TMS baseline measurement; pain intensity average was above four at the training blocks- | A modified version of the sequential visual isometric pinch task in three phases (baseline 1, baseline 2, and acquisition) | Movement time, accuracy, and a skill measure | No sig effect of cutaneous pain on motor skill acquisition |

| measuring by NPRS |  |  |  |  |  |
| --- | --- | --- | --- | --- | --- |
| Dancey et al (2014) | Healthy (N=24) (pain group N=12, f=7, age: 24.5 ± 6.6 yrs); (control group N=12, f=6, age: 23.4 ± 2 yrs) | Capsaicin gel for pain group and topical cream for control group in the lateral part of the right elbow; pain intensity average was 5 in the post-application phase- measuring by NPRS | A repetitive typing task applying the middle three fingers | Motor training accuracy; reaction time | An improvement in motor performance in the presence of tonic pain |
| Ingham et al (2011) | Healthy (N=9, m=7, age: 24 ± 1.1) | Hypertonic saline for pain group; remote pain in the infrapatellar fat pad of the knee and local pain in the FDI; pain intensity average was 0.2 ± 0.4 (vehicle control), 1.7 ± 1 (FDI pain) and 2.1 ± 1.6 (remote pain)- measuring by NRS | A quick movements of finger (the right index finger) adduction | Training performance based on peak acceleration of index finger movement | No sig effect of pain on motor performance |
| Salomoni et al (2019) | Healthy (N=22) (pain group N=11, f=7, age: 24.5 ± 6.6 yrs); (control group N=11, f=5, age: 23.4 ± 2 yrs) | Hypertonic saline to the anterior deltoid muscle prior to baseline 2 and before the force field 1 in the first day; the intensity of pain was 4.2 ± 0.3 (out of 10) in the first injection and 3 ± 0.4 in the second injection- measuring by NPRS | A reaching task in the presence of force field adaptation in six phases (baseline1, baseline2, force field 1, force field 2, washout 1, force field 2 and washout 2) | Movement accuracy based on the peak perpendicular error and the peak hand velocity; muscle activity of anterior and posterior deltoid, biceps brachii, triceps brachii, and pectoralis major | No sig effect of pain on the final performance; experimental pain group used different strategies to perform the same task compared to the control group |
| Boudreau et al (2007) | Healthy (N=9, m=7, age: 24 ± 1.1) | Capsaicin gel for pain group and vehicle cream for the control group to the tongue; pain intensity average was between 4 and 6 during the task- measuring by VAS | A tongue-protrusion task | A performance score based on the time that participants kept the cursor within the target box | A sig and negative effect of pain on the overall performance score |
| Boudreau et al (2010) | Healthy (N=26) (lidocaine group N=9, f=6, age: 24.6 ± 1.1 yrs); (control group N=9, f=3, age: 24 ± 3.5 yrs); (capsaicin group N=8 f=2, age: 23 ± 2.5 yrs) | Capsaicin and lidocaine gels for pain groups and vehicle cream for the control group to the dorsum of the tongue before the first task; pain intensity was maintained above 4- measuring by NRS | A tongue-protrusion task | (Overall, a tongue-task trial, initial, within-session gains) motor performance | A sig decrease in motor performance in the pain groups than the control group |
| Gallina et al (2018) | Healthy (N=14, f=7, age: 18-47) | Hypertonic saline to the infrapatellar fat pad, distal vastus medialis, proximal vastus medialis, and vastus lateralis; the intensity of pain was approximately 3 in four different pain locations- measuring by NRS | Isometric knee extension contraction | Muscle activation of VMO, VL, and RF; isometric knee extension force | A sig alteration in both muscle activation and knee extension force in response to different locations of pain |

Abbreviations: f female, m male, yrs years, VMO vastus medialis oblique, VL vastus lateralis, RF rectus femoris, sig significant, VAS visual analogue scale, NRS numerical rating scale, TMS transcranial magnetic stimulation, FDI first dorsal interosseous

### Methodological quality

The methodological quality of the included studies was assessed based on the modified version of Downs and Black checklist, which is provided in Table 2. Out of 17 studies, eleven studies (Bilodeau et al., 2016; Boudreau et al., 2010; Bouffard et al., 2014, 2016; Dancey et al., 2016a; Dancey et al., 2014; Dancey et al., 2016b; Dancey et al., 2018, 2019; Ingham et al., 2011; Salomoni et al., 2019) were evaluated as fair quality and 6 articles as poor quality (Arieh et al., 2021; Boudreau et al., 2007; Bouffard et al., 2018; Gallina et al., 2018; Lamothe et al., 2014; Mavromatis et al., 2017). Inter-rater reliability was 0.72 between the assessors who evaluated the methodological quality of the included articles.

**Table 2.**
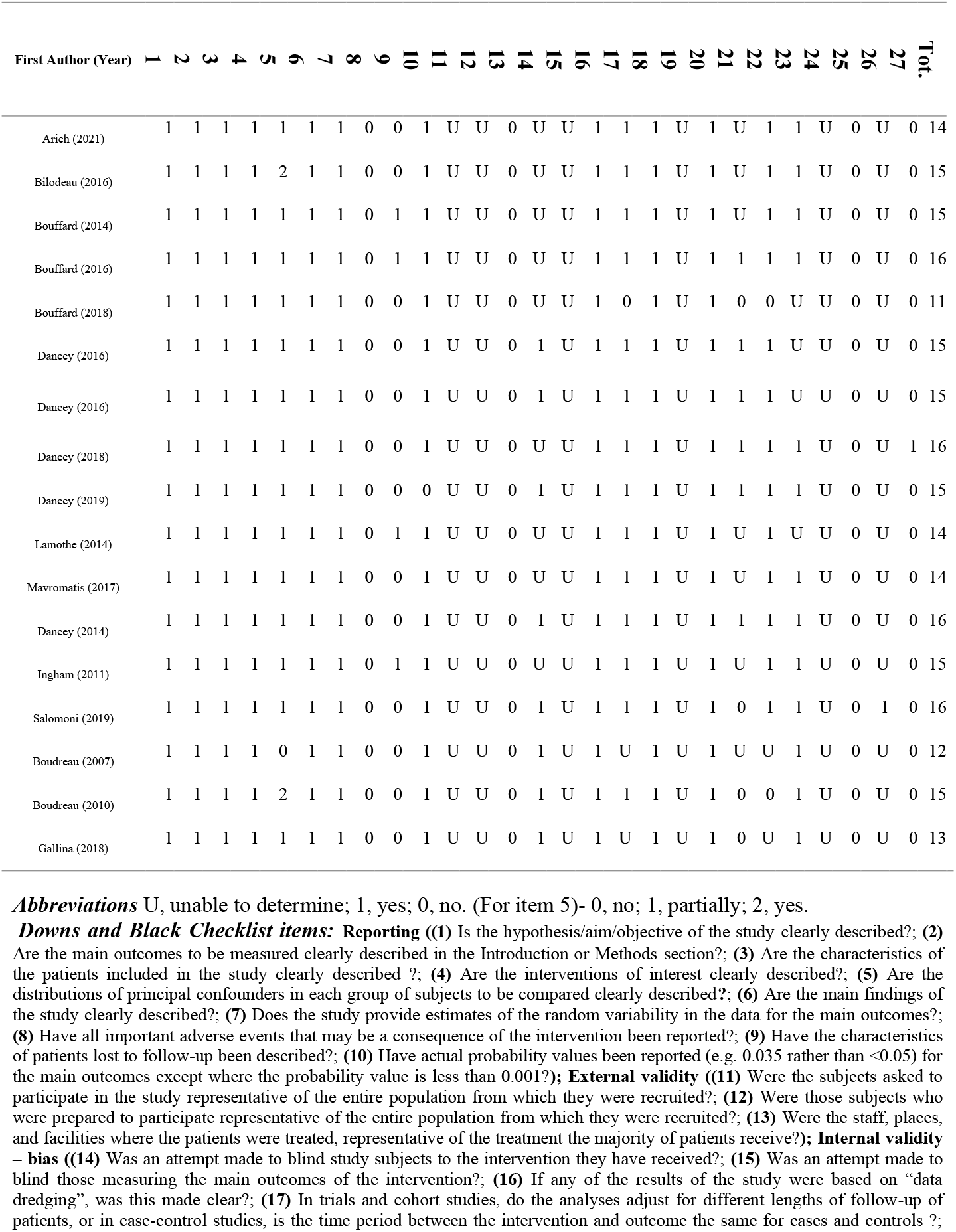
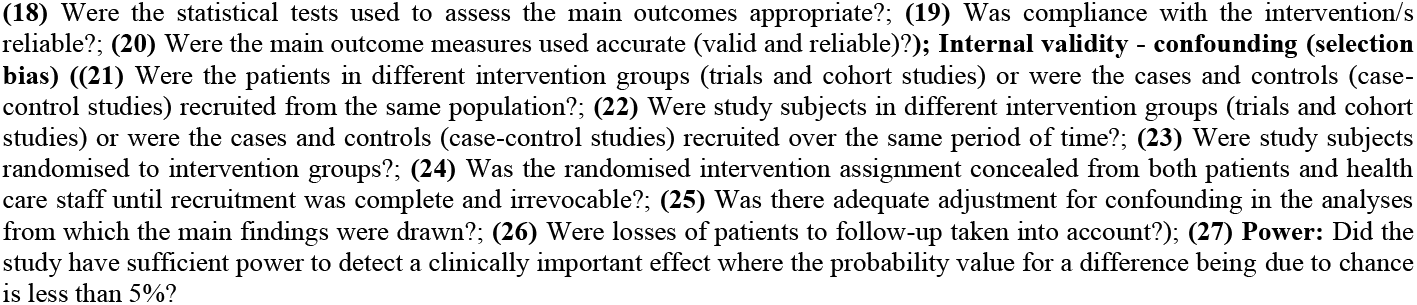
Quality Assessment of the Included Studies

| First Author (Year) | 1 | 2 | 3 | 4 | 5 | 6 | 7 | 8 | 9 | 10 | 11 | 12 | 13 | 14 | 15 | 16 | 17 | 18 | 19 | 20 | 21 | 22 | 23 | 24 | 25 | 26 | 27 | Tot. |
| --- | --- | --- | --- | --- | --- | --- | --- | --- | --- | --- | --- | --- | --- | --- | --- | --- | --- | --- | --- | --- | --- | --- | --- | --- | --- | --- | --- | --- |
| Arieh (2021) | 1 | 1 | 1 | 1 | 1 | 1 | 1 | 0 | 0 | 1 | U | U | 0 | U | U | 1 | 1 | 1 | U | 1 | U | 1 | 1 | U | 0 | U | 0 | 14 |
| Bilodeau (2016) | 1 | 1 | 1 | 1 | 2 | 1 | 1 | 0 | 0 | 1 | U | U | 0 | U | U | 1 | 1 | 1 | U | 1 | U | 1 | 1 | U | 0 | U | 0 | 15 |
| Bouffard (2014) | 1 | 1 | 1 | 1 | 1 | 1 | 1 | 0 | 1 | 1 | U | U | 0 | U | U | 1 | 1 | 1 | U | 1 | U | 1 | 1 | U | 0 | U | 0 | 15 |
| Bouffard (2016) | 1 | 1 | 1 | 1 | 1 | 1 | 1 | 0 | 1 | 1 | U | U | 0 | U | U | 1 | 1 | 1 | U | 1 | 1 | 1 | 1 | U | 0 | U | 0 | 16 |
| Bouffard (2018) | 1 | 1 | 1 | 1 | 1 | 1 | 1 | 0 | 0 | 1 | U | U | 0 | U | U | 1 | 0 | 1 | U | 1 | 0 | 0 | U | U | 0 | U | 0 | 11 |
| Dancey (2016) | 1 | 1 | 1 | 1 | 1 | 1 | 1 | 0 | 0 | 1 | U | U | 0 | 1 | U | 1 | 1 | 1 | U | 1 | 1 | 1 | U | U | 0 | U | 0 | 15 |
| Dancey (2016) | 1 | 1 | 1 | 1 | 1 | 1 | 1 | 0 | 0 | 1 | U | U | 0 | 1 | U | 1 | 1 | 1 | U | 1 | 1 | 1 | U | U | 0 | U | 0 | 15 |
| Dancey (2018) | 1 | 1 | 1 | 1 | 1 | 1 | 1 | 0 | 0 | 1 | U | U | 0 | U | U | 1 | 1 | 1 | U | 1 | 1 | 1 | 1 | U | 0 | U | 1 | 16 |
| Dancey (2019) | 1 | 1 | 1 | 1 | 1 | 1 | 1 | 0 | 0 | 0 | U | U | 0 | 1 | U | 1 | 1 | 1 | U | 1 | 1 | 1 | 1 | U | 0 | U | 0 | 15 |
| Lamothe (2014) | 1 | 1 | 1 | 1 | 1 | 1 | 1 | 0 | 1 | 1 | U | U | 0 | U | U | 1 | 1 | 1 | U | 1 | U | 1 | U | U | 0 | U | 0 | 14 |
| Mavromatis (2017) | 1 | 1 | 1 | 1 | 1 | 1 | 1 | 0 | 0 | 1 | U | U | 0 | U | U | 1 | 1 | 1 | U | 1 | U | 1 | 1 | U | 0 | U | 0 | 14 |
| Dancey (2014) | 1 | 1 | 1 | 1 | 1 | 1 | 1 | 0 | 0 | 1 | U | U | 0 | 1 | U | 1 | 1 | 1 | U | 1 | 1 | 1 | 1 | U | 0 | U | 0 | 16 |
| Ingham (2011) | 1 | 1 | 1 | 1 | 1 | 1 | 1 | 0 | 1 | 1 | U | U | 0 | U | U | 1 | 1 | 1 | U | 1 | U | 1 | 1 | U | 0 | U | 0 | 15 |
| Salomoni (2019) | 1 | 1 | 1 | 1 | 1 | 1 | 1 | 0 | 0 | 1 | U | U | 0 | 1 | U | 1 | 1 | 1 | U | 1 | 0 | 1 | 1 | U | 0 | 1 | 0 | 16 |
| Boudreau (2007) | 1 | 1 | 1 | 1 | 0 | 1 | 1 | 0 | 0 | 1 | U | U | 0 | 1 | U | 1 | U | 1 | U | 1 | U | U | 1 | U | 0 | U | 0 | 12 |
| Boudreau (2010) | 1 | 1 | 1 | 1 | 2 | 1 | 1 | 0 | 0 | 1 | U | U | 0 | 1 | U | 1 | 1 | 1 | U | 1 | 0 | 0 | 1 | U | 0 | U | 0 | 15 |
| Gallina (2018) | 1 | 1 | 1 | 1 | 1 | 1 | 1 | 0 | 0 | 1 | U | U | 0 | 1 | U | 1 | U | 1 | U | 1 | 0 | U | 1 | U | 0 | U | 0 | 13 |

**Abbreviations** U, unable to determine; 1, yes; 0, no. (For item 5)- 0, no; 1, partially; 2, yes.

### Cutaneous pain

Thirteen studies (Arieh et al., 2021; Bilodeau et al., 2016; Boudreau et al., 2010; Boudreau et al., 2007; Bouffard et al., 2014, 2016; Dancey et al., 2016a; Dancey et al., 2014; Dancey et al., 2016b; Dancey et al., 2018, 2019; Lamothe et al., 2014; Mavromatis et al., 2017) applied capsaicin gel, resulting in cutaneous pain, to understand the effect of acute pain on motor learning. There was no consensus surrounding the effect of cutaneous pain on motor learning among these studies. Specifically five studies (Arieh et al., 2021; Bilodeau et al., 2016; Bouffard et al., 2016; Dancey et al., 2018; Mavromatis et al., 2017) reported no significant change in motor learning in response to acute pain; but eight studies (Boudreau et al., 2010; Boudreau et al., 2007; Bouffard et al., 2014; Dancey et al., 2016a; Dancey et al., 2014; Dancey et al., 2016b; Dancey et al., 2019; Lamothe et al., 2014) demonstrated a meaningful influence of cutaneous pain on motor learning. Each of the studies measured different motor learning variables applied during different motor tasks to study the effect of pain. In this context, Dancey and colleagues carried out a series of studies (Dancey et al., 2016a; Dancey et al., 2014; Dancey et al., 2016b; Dancey et al., 2018, 2019) to reveal the role of cutaneous pain on motor learning during typing and tracing series of sinusoidal patterns. The results of their studies show a meaningful positive influence of the experimental pain on skill acquisition learning (Dancey et al., 2016a; Dancey et al., 2014; Dancey et al., 2016b; Dancey et al., 2019) and retention (Dancey et al., 2016b; Dancey et al., 2019). While one study demonstrated that local pain improved retention of learning compared to remote pain (Dancey et al., 2016b), another study did not observe any significant effect on motor learning variables in response to different pain locations (remote, local, or contralateral) (Dancey et al., 2018). Two studies that used finger-tapping (Bilodeau et al., 2016) and sequential visual isometric pinch (Mavromatis et al., 2017) tasks to show the effect of cutaneous pain on motor learning found no significant differences in the pain group compared to the control group for both motor learning acquisition (Bilodeau et al., 2016; Mavromatis et al., 2017) and retention (Bilodeau et al., 2016). Boudreau and colleagues (Boudreau et al., 2010; Boudreau et al., 2007) demonstrated a significant negative influence of experimental cutaneous pain on overall motor performance during a tongue-protrusion task. In addition, while Lamothe et al (2014) indicated a significant improvement in motor performance in response to cutaneous pain during a new reaching adaptation task, the pain group demonstrated a larger final error to perform the same task compared to the control group in both acquisition and retention phases (Lamothe et al., 2014). Bouffard et al (2014) revealed a significant decrease in the performance during the retention test of motor in the experimental cutaneous pain group during a novel locomotor adaptation task, with no difference between the groups during the initial learning (Bouffard et al., 2014). In a related study (Bouffard et al., 2016), they did not observe any considerable difference in either the learning or the retention of the new locomotor task, but in this study the capsaicin gel was applied in both the learning and retention tests. Finally, Arieh et al. (2021) demonstrated no significant difference in movement accuracy in both acquisition and retention phases of motor learning in response to the experimental pain during dart-throwing skill (Arieh et al., 2021).

### Muscle pain

Four studies (Bouffard et al., 2018; Gallina et al., 2018; Ingham et al., 2011; Salomoni et al., 2019) evaluated motor learning by injecting hypertonic saline, resulting in muscle pain, during different tasks. All studies revealed no significant effect of experimental muscle pain on motor performance. Specifically, Bouffard and colleagues (2018) did not observe any meaningful change in acquisition and retention of a novel locomotor adaptation task in response to the experimental pain (Bouffard et al., 2018). Ingham et al. (2011) also demonstrated no significant effect on motor learning in a finger adduction task in which muscle pain was applied in different locations. While Salomoni and colleagues (2019) did not observe any meaningful alteration in final motor performance in the pain group compared to the control group during a reaching adaptation task, those who experienced the experimental muscle pain applied a distinct strategy to perform the task in comparison with the control group (Salomoni et al., 2019). The experimental group also produced the same strategy in the next day in the absence of pain. A similar result was found by Gallina and colleagues in which the muscle pain location produced lasting changes in the muscle activation pattern during an isometric knee extension contraction task (Gallina et al., 2018).

## Discussion

The aim of the current study was to understand the effect of acute pain on motor learning among healthy individuals. Inconsistent results have been reported surrounding this topic in the literature; however, most of these studies are in agreement with the negative consequences of acute pain in the learning process. Moreover, while some studies did not demonstrate any significant effect of experimental pain on skill learning acquisition and retention, they indicated those who experienced pain produced a distinct strategy to perform the novel task compared to control groups such that the participants displayed the same strategy in pain resolution even after one week.

Learning new movement patterns is an integral part of sport and rehabilitation (Franklin & Wolpert, 2011), while pain can give rise to alterations in the learning process. Results of the studies that have examined the effect of experimental pain on motor learning corroborate the role of acute pain on changes in motor adaptation; however, the studies demonstrated contradictory findings. Specifically, a series of investigations by Dancey and colleagues (Dancey et al., 2016a; Dancey et al., 2014; Dancey et al., 2016b; Dancey et al., 2019) revealed a positive and meaningful effect of cutaneous pain on the learning process. It has been suggested that pain can lead to an increase in attention while performing a dynamic task thereby those who experience pain can execute a function with lesser errors than no pain condition (Dancey et al., 2016b; Hazeltine et al., 1997). In this context, Dancey and colleagues reported an improvement in skill learning acquisition and retention in the presence of experimental pain because of attention mechanism, in which local pain brought about a better overall motor performance compared to remote pain (Dancey et al., 2016a; Dancey et al., 2014; Dancey et al., 2016b; Dancey et al., 2018). It was argued that local pain may result in more attention to the part of the body underlying learning (Dancey et al., 2016b), which in turn can lead to more changes in cortical neuroplasticity (Rosenkranz & Rothwell, 2004), and subsequently improve in the learning of motor tasks. A recent study (Tumialis et al., 2020) indicated that external attention can engender more accuracy and better performance in comparison to internal attention, including performing a task in the presence of pain. Whereas Mavromatis and colleagues (Mavromatis et al., 2017) applied a similar experimental pain model compared to the work of Dancey (Dancey et al., 2016a; Dancey et al., 2014; Dancey et al., 2016b; Dancey et al., 2018), they did not observe any significant effect of acute pain on the skill learning acquisition. However, the training-related alterations in corticospinal excitability were showed a similar result to Dancey’s study in the presence of cutaneous pain (Mavromatis et al., 2017). In contrast to the previous studies, Boudreau and colleagues (Boudreau et al., 2010; Boudreau et al., 2007) reported a significant negative influence of cutaneous pain on overall performance scores. While all of these studies applied a similar experimental tonic pain, the studies used a range of tasks to understand the effect of pain on motor learning which might explain some of the differences in the findings. In particular, different motor tasks and different types of learning depend on different brain mechanisms and brain areas (Doya, 2000; Ghilardi et al., 2000). These differences may actually be one of the major reasons why we find inconsistent results of the effect of pain on motor learning (Bilodeau et al., 2016). For example, force field adaptation and sequence learning tasks can rely on cortico-cerebellar and cortico-striatal plasticity, respectively (Doyon & Benali, 2005). Similarly, Seidler and colleagues found large differences in brain activation even within a similar task; where performance with smaller errors during movements to large easy to reach targets were associated with greater activation in the contralateral primary motor cortex, premotor cortex and the basal ganglia, and larger errors during movements to small targets associated with greater activation in the ipsilateral motor cortex, insular cortex, cingulate motor area, and multiple cerebellar regions (Seidler et al., 2004). That is, variations in the task difficulty (e.g. target size) can influence the degree of feedforward relative to feedback control that contributes to the task performance, and therefore the specific brain areas involved. It is very likely that the different circuitry and adaptation mechanisms involved in different motor tasks have different reactions to painful stimuli.

While the studies that applied cutaneous pain to evaluate motor learning revealed contradictory results, most research (Bouffard et al., 2018; Ingham et al., 2011; Salomoni et al., 2019) indicated no significant effect of experimental muscle pain on motor learning outcomes. Indeed, it has been reported that these two experimental pain models interact distinctly with neural processes that are responsible for motor adaptation (Henderson et al., 2006), which in turn can lead to observe different results in skill learning acquisition and retention in response to cutaneous or muscle pain models. This distinction between cutaneous and muscle pain models was particularly clear in a series of studies by Bouffard and colleagues (Bouffard et al., 2014, 2016; Bouffard et al., 2018) demonstrating motor learning outcomes in response to experimental muscle and cutaneous pain models during a locomotor adaptation task. Specifically, there was no alteration in skill learning acquisition or retention in the presence of experimental muscle pain (Bouffard et al., 2018). However they did find a meaningful reduction in retention (but not acquisition) of the same test in the presence of the cutaneous pain model (Bouffard et al., 2014). However, a follow-up study showed that cutaneous pain had no effect on either the acquisition or the retention as long as this pain was also applied during the test for retention (Bouffard et al., 2016). That is, it appeared that the cutaneous pain acted as a contextual signal for the selection of the newly learned locomotion model (Bouffard et al., 2016), similar to the manner that visual, proprioceptive and vestibular signals can be used to learn and recall different motor memories (Howard & Franklin, 2016; Howard et al., 2012; Sarwary et al., 2015). Although no considerable influence of cutaneous pain on motor performance or motor learning was shown, Bouffard and colleagues reported that participants in the pain group produced a distinct strategy compared to the control group to perform a locomotion task. Specifically, they found that participants had a different pattern of kinematic errors in the presence of pain during walking suggesting the pain group used less predictive compensation (anticipatory strategies) for the changes in the task (Bouffard et al., 2016). This finding was supported by several other studies (Arieh et al., 2021; Bouffard et al., 2018; Salomoni et al., 2019) which found participants exposed to experimental pain expressed a different strategy for motor adaptation compared to control participants despite no significant change in overall motor performance in the skill learning acquisition and retention. Notably, Salomoni et al. (2019) found that participants who experienced experimental muscle pain produced less co-contraction and muscle activation of the elbow and shoulder muscles compared to the pain-free control group during a reaching task, and that this distinct motor strategy was continued on the next day (retention) despite the pain no longer being present. This smaller muscle co-contraction could potentially reduce joint stability in the coordination of musculoskeletal system (Franklin et al., 2007; Franklin et al., 2013) and subsequently increase the potential for musculoskeletal injuries during sport training and rehabilitation (Henriksen et al., 2007). Arieh and colleagues also showed a similar motor performance in response to experimental pain compared to the control group during dart-throwing skill; however, participants in the pain group showed different coordination patterns in the shoulder-elbow and elbow-wrist joints to perform the task even one week later. These different movement patterns may be a strategy to decrease pain while still performing the motor adaptation task, as suggested by Hodges and Tucker (Hodges & Tucker, 2011), in which pain affects the redistribution of activity within and between muscles (Muceli et al., 2014; van den Hoorn et al., 2015) to perform a motor task with a pain-free movement pattern. While this mechanism might be used to reduce pain during the learning process, such an alteration could potentially be associated with repercussions for the health condition of joints over longer time periods. That is, redistribution of muscle function can bring about changes in natural biomechanics of the joints by increasing joint load (Hodges & Tucker, 2011). These changed patterns of muscle activation or joint coordination can then persist over long periods of time either due to use-dependency (Burdet et al., 2013; Diedrichsen et al., 2010; Doya, 2000) or because the adaptation process resulted in local minimum of the solution space (Burdet et al., 2013).

The results of the present systematic review need to be interpreted with the consideration of the following methodological issues. First, sleep between the acquisition and retention phases can be a factor that also influences motor learning, and only one study (Bilodeau et al., 2016) considered this issue before evaluating motor learning in response to experimental pain. Second, as physical and mental performances can fluctuate due to circadian rhythm; it has been suggested that physical and mental tests should be measured at the same time of day, especially for studies that apply repeated measurement protocols (Vitale & Weydahl, 2017). None of the studies reported this possible factor when assessing motor learning in response to experimental pain models. Third, the difficulty of a new motor adaptation task can result in a challenge to the success of performing a task (Guadagnoli & Lee, 2004); hence, the optimal challenge point should be determined when designing a motor learning task, to ensure that sufficient outcome measurement sensitivity is obtained. Otherwise, if the tasks are too difficult, too simple, or the performance measurement is too imprecise, a study may find no difference between control and pain groups even when a difference actually exists, producing a type 2 error (false negative). None of the included studies mentioned this important issue. In addition, only one study (Dancey et al., 2018) reported an adequate sample size for carrying out its research. Finally, the studies included in this review particularly focused on the effect of pain on motor learning in young healthy individuals, so further studies are needed to verify if similar effects are found in children and older adults, as well as expanding to patients and chronic pain conditions.

## Conclusion

Overall, this systematic review found heterogeneous results regarding experimental pain models’ influence on motor learning. In particular, although experimental pain models have been reported to lead to changes in the skill learning acquisition and retention, many studies have also shown unaltered adaptation in motor learning outcomes. Finally, several studies have shown that distinct strategies have been observed in the pain group even after pain resolution. These variable results highlight the need for further studies to clarify the effect of pain on motor learning.

## Data Availability

The present study is a systematic review wherein there is no data regarding experimental design.

## Notes

### Competing Interest Statement

The authors have declared no competing interest.

### Funding Statement

This research did not receive any specific grant from funding agencies in the public, commercial, or not-for-profit sectors.

### Author Declarations

The current study is a systematic review in which no experiment was carried out whereby it did not require any ethical approval.

